# A novel quantitative method via the degree of transverse colon ptosis for chronic constipation assessment

**DOI:** 10.1101/2020.04.09.20059527

**Authors:** Kongliang Sun, Qun Qian, Jinxiang Hu, Weicheng Liu, Yuntian Hong, Wenwen Zhang, Hui-Xuan Xie, Bo Liu, Xianghai Ren, Changlei Xi, Hong Yan, Congqing Jiang, Xiaoyu Xie

## Abstract

**BACKGROUND:** Assessment of colonic transit tend to be more subjective and qualitative. This study aimed to evaluate the capability of our new quantitative scale to predict the subtypes of constipation and assess symptom severity of patients with slow transit constipation.

**METHODS:** A retrospective cohort population was assembled, consisting of adult patients with chronic constipation who underwent both colonic transit test and defecography between 2012 and 2019. Radiological parameters were measured on AXRs. The Luojia score was introduced to convey the vertical distance from the splenic flexure to the lowest point of the transverse colon, representing the degree of transverse colon ptosis. Patients with slow transit constipation only were especially required to complete the Wexner Constipation Scale (WCS) and Hospital Anxiety and Depression Scale (HADS) for clinical severity assessment.

**FINDINGS:** Of 368 patients, 191 patients (51·9%) showed slow colonic transit, and patients with slow colonic transit were more likely to have severe ptosis of the transverse colon on AXRs. Patients with slow colonic transit had a significantly higher Luojia score than those with normal colonic transit (p˂0·001). A cut-off of 195 mm was used to distinguish slow colonic transit. A significant difference in Luojia score was also found between patients with obstructed defecation syndrome and normal patients, and a cut-off of 140 mm was identified. In patients with slow transit constipation, there was a strong correlation between Luojia score and WCS (r=0·618) and a moderate correlation between Luojia score and HADS-Anxiety (r=0·507). These results indicated that the Luojia score is a reliable predictor of symptom severity and psychological condition in patients with slow transit constipation.

**INTERPRETATION:** The Luojia score might be a new quantitative, precise method in the assessment of constipation.

**FUNDING:** The National Natural Science Foundation of China and the Clinical Research Special Fund of Wu Jieping Medical Foundation.

**Research in context:** *Evidence before this study:* We searched PubMed for papers published between Feb 1, 2000, and Jan 1, 2019, with the keywords “transverse colonic ptosis” OR “abdominal x-ray” AND “constipation” OR “colonic transit”. No restrictions on study type or language were implemented. Our search retrieved studies on the use of stool burden score on AXR in the assessment of constipation but no studies to use transverse colonic ptosis to evaluate colonic transit.

*Added value of this study:* We established a Luojia score which was defined as the vertical distance from the splenic flexure to the lowest point of transverse colon on the abdominal x-ray (AXR) that representing the degree of transverse colon ptosis. A retrospective cohort study of 368 patients proved that Luojia score has high sensitivity and specificity in distinguishing slow colonic transit and normal colonic transit as well as obstructed defecation syndrome and normal group. We were satisfied to found that in patients with slow transit constipation, there was a strong correlation between Luojia score and WCS (r=0·618) and a mediate correlation between Luojia score and HADS-A (r=0·507).

*Implications of all the available evidence:* Precise assessment and evaluation of colonic transit play an important role in clinical diagnosis and treatment of constipation patients. Our result proved that Luojia Score is a simple and effective assessment system of certain clinic value in in identifying patients with constipation and is a potential predictor of symptom severity.

## Introduction

Chronic constipation is a common gastrointestinal disorder worldwide, with a prevalence of 16%^1^ in Western countries and a similar prevalence of 14–18% in China.^2,3^ It can be caused by slow transit of chyme within the colon or impaired rectal evacuation.^4^ Long-term constipation can induce abdominal distension, abdominal pain, defecation difficulty, and weakened or even absent sense of defecation.

Colonic transit is mainly evaluated via radiopaque markers on plain film abdominal X-ray (AXR). This is an inexpensive visualization method that is widely applied in clinical settings and plays important roles in distinguishing constipation subgroups, such as normal or slow transit constipation (STC).^5^ Despite the frequency with which AXR is applied, studies have suggested that it has many drawbacks because of radiation exposure of patients, requirement for multiple ambulatory visits, and dependence on experience of radiologists.^6^ Additionally, there is no ‘gold standard’ method for this test.^7^

Although gamma scintigraphy, magnetic tracking system approach, and magnetic resonance imaging (MRI) provided new imaging detection methods in the identification of colonic transit disorder and stool burden, has been considered as a reliable alternative radiopaque marker method in the assessment of colonic transit, due to the limitation of these methods, a concise and efficient assessment is needed for constipation evaluation.^8,9^

Transverse colon ptosis is common in patients with chronic constipation, which may be the result of long-term constipation. However, more importantly, it may also cause difficulty in pushing stool in the colon and aggravate constipation.^10-13^ To date, the assessment of transverse colon ptosis has not yet gained particular attention. The lack of a well-validated, standardized assessment method of constipation severity has been a significant obstacle in treatment strategy decisions.

This study aimed to investigate the relationship between the degree of transverse colon ptosis on AXRs and severity of constipation. Evidence linking transverse colon ptosis and disease severity could provide an objective means of identifying patients with treatment-refractory constipation earlier in their evaluation.

## Methods

### Subjects

We assembled a retrospective cohort population consisting of adult patients (aged > 18 years) with chronic functional constipation that were diagnosed according to the Rome IV criteria. The patients were divided into three groups based on colonoscopy results, colonic transit time (CTT), and defecography findings described in our previous study: STC, obstructed defecation syndrome (ODS), and mixed constipation (STC+ODS).^14^ Patients were excluded if they fulfilled any of the following criteria: (1) previous abdominal and pelvic radiotherapy; (2) previous abdominal and anal surgery that may affect the patient’s bowel function (spinal, gynaecological, obstetric, or abdominal surgery and pelvic floor reconstruction or repair); (3) pelvic-floor dyssynergia; (4) history of malignancy; (5) megacolon, strictures, intestinal obstruction, and other anatomical abnormalities responsible for constipation; and (6) chronic constipation due to a secondary cause (medications, diabetes mellitus, hypothyroidism, etc.). Patient data were recruited from outpatient inquiry and hospital information system. The patient demographics (i.e., age of first constipation visit, sex, weight, and height) were collected as the characterization of patient population. This study was approved by the medical ethics committee of Zhongnan Hospital of Wuhan University.

### Radiographic examinations

The barium suspension method was applied for CTT to better investigate the anatomical abnormality and locations of each segments of the whole colon.^5^ Patients were prohibited from using laxatives, enemas, or any kind of suppositories for at least 3 days before the study. They were required to keep the usual diet and activities during the examination. Patients swallowed up to 50 mL of 200% (w/v) barium sulphate suspension as soon as possible on the first day, and AXRs in a standing position were obtained at 6 h, 24 h, 48 h, and 72 h after ingestion to record the distribution and discharge of barium suspension in the colon. CTT is determined according to the previous criteria.^5^ Defecography or magnetic resonance defecography was performed in patients with ODS symptoms. ^15^

### Radiological parameters

Visible radiopaque suspension was easily identified on AXRs with bony landmarks and the outlines of the colon, which was helpful in precisely measuring the following radiological parameters: The vertical distances from the hepatic flexure to the right iliac crest (HI), from the splenic flexure to the left iliac crest (SI), from the lowest point of transverse colon to the L4-L5 interspace (of the same height to the attachment of both sides of the iliac crest^16^) (TI), from the costophrenic angle to the splenic flexure (CS), and from the splenic flexure to the hepatic flexure (SH) were measured on the AXRs. Meanwhile, the Luojia score was a new concept presented in this study. It is defined as the vertical distance from the splenic flexure to the lowest point of the transverse colon (SI minus TI), which represents the degree of transverse colon ptosis.

### Assessment of symptom severity and psychological condition

Constipation symptom severity was measured using the Wexner Constipation Scale (WCS),^17^ an eight-term measure of constipation severity scores ranging from 0 to 30, with 0 indicating normal and 30 indicating severe constipation. Similarly, the Hospital Anxiety and Depression Scale (HADS), a 14-term measure for the detection of anxiety and depression, was applied to determine the psychological condition.^18^ Scores ≥8/21 for both depression and anxiety are considered diagnostic with a sensitivity and specificity of approximately 0·80.^19^ For precise assessment of symptom severity and psychological condition, patients with STC only were especially required to complete the WCS and HADS.

### Statistical analysis

For continuous variables, normality of distribution was tested using the Kolmogorov-Smirnov test. Data were presented as mean ± standard deviation for variables with normal distribution and median (interquartile range 25, 75) for variables with abnormal distribution. Comparisons among continuous independent variables were performed using t-tests or Mann-Whitney U tests. Comparisons among categorical variables were performed using the chi-square tests. In terms of three or more groups, comparisons of median values were performed using the Kruskal-Wallis H test with post hoc analysis. Correlation between radiological parameters and clinical features was determined using Pearson correlation coefficient. For the absolute value of r, 0–0·19 is regarded as extremely weak, 0·2–0·39 as weak, 0·40–0·59 as moderate, 0·6–0·79 as strong, and 0·8–1·0 as extremely strong correlation. A p-value <0·05 indicated statistical significance. Receiver operating characteristic (ROC) curves were created, and an area under the ROC curve (AUC) was calculated. Then, Youden’s J statistic was used to determine an optimal cut-off point (maximizing both sensitivity and specificity) for the Luojia score. Statistical processing was conducted using SPSS Statistics version 24·0.

## Results

### Cohort characteristics

A total of 368 patients were included in this cohort study, 278 (77·5%) of them were women, and the mean age was 55·22±13·16 years. Among them, 191 patients (51·9%) were identified to have slow colonic transit by colonic transit test, and women are more likely to have slow colonic transit (80·6% vs 70·1%, P=0·018). There was no significant difference in age between the slow colonic transit group and normal colonic transit group (55·93±11·83 vs 54·45±14·44 years, p=0·280). Although the two groups of patients were of the same height (1·62±0·06 vs 1·62±0·07 m, p=0·255), the slow colonic transit group seemed to have lower body weight (58·56±7·99 vs 63·44±8·46 kg, p˂0·001) and lower body mass index (BMI) (22·43±3·15 vs 24·08±3·20 kg/m^2^, p˂0·001). The slow colonic transit group had longer intervals of bowel movement (6·89±2·38 vs 2·42±2·08 days, p˂0·001) and spent more time for each attempt of evacuation (27·23±12·20 vs 12·71±12·47 min, p˂0·001).

For precise assessment of colonic transit and better description of constipation symptom, we categorized the patients into four subgroups: 52 patients (14·1%) had STC+ODS, 139 patients (37·8%) had STC, 52 patients (14·1%) had ODS, and the remaining 125 patients (34·0%) did not have STC or ODS (Figure 1). The symptoms claimed by the patients differed from each subgroup of colonic transit, and adjusted residuals were below the observed frequency (Table 2). Patients with slow colonic transit (including STC and STC+ODS) more commonly had the complaint of absent sense of defecation (p˂0·001) and dependence on laxatives or enemas (p=0·020) than patients with ODS. Nevertheless, patients with ODS were more likely to have multiple attempts to defecate (p˂0·001) and incomplete evacuation (p=0·034). Additionally, there was no significant difference in the complaint of abdominal pain (p=0·195) and abdominal distension (p=0·568) in the three groups of patients (Table 2).

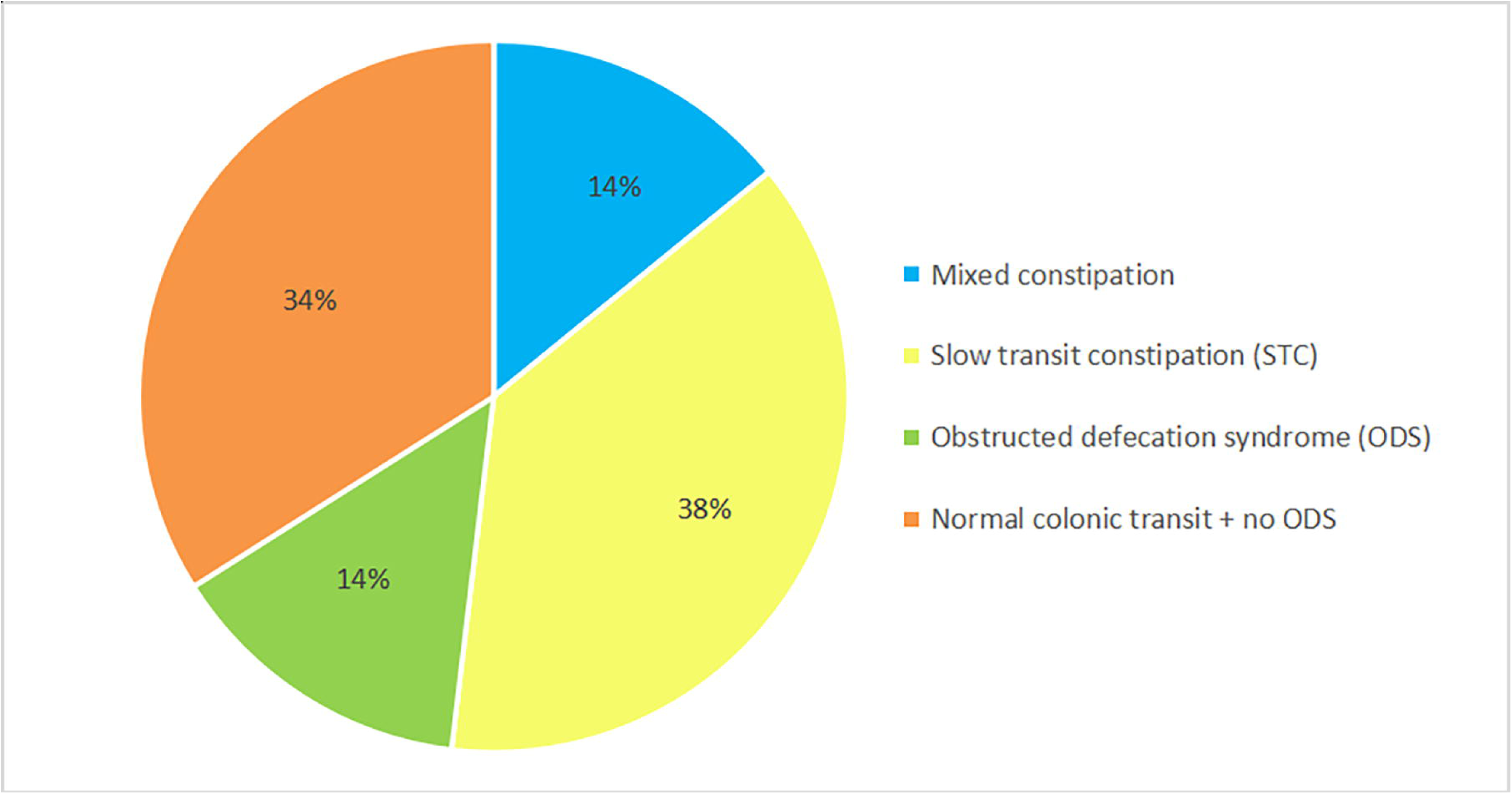

**Table 1:** Cohort characteristics

|  | Total<br>(N=368) | Normal colonic<br>transit<br>(N=177) | Slow colonic transit<br>(N=191) | p value |
| --- | --- | --- | --- | --- |
| Age(yrs) | 55.21±13.16 | 54.45±14.44 | 55.93±11.83 | 0.280 |
| Female sex,N(%) | 277(75.3) | 124(70.1) | 154(80.6) | 0.018 |
| Height(m) | 1.62±0.06 | 1.62±0.07 | 1.62±0.06 | 0.255 |
| Weight(Kg) | 60.90±8.57 | 63.44±8.46 | 58.56±7.99 | <0.001 |
| BMI(kg/m<sup>2</sup>) | 23.22±3.27 | 24.08±3.20 | 22.43±3.15 | <0.001 |
| Interval of bowel<br>movement(days) | 4.74±3.17 | 2.42±2.08 | 6.89±2.38 | <0.001 |
| Length of time per<br>attempt(mins) | 20.24±14.30 | 12.71±12.47 | 27.23±12.20 | <0.001 |
| HI(mm) | 97.21±42.62 | 113.95±43.59 | 81.69±35.28 | <0.001 |
| SI(mm) | 143.55±38.19 | 146.57±40.44 | 140.75±35.86 | 0.144 |
| TI(mm) | -38.38±86.32 | 28.69±72.23 | -100.55±38.52 | <0.001 |
| CS(mm) | 29.91±31.92 | 37.94±32.27 | 22.46±29.79 | <0.001 |
| SH(mm) | 46.34±37.52 | 32.62±32.15 | 59.06±37.73 | <0.001 |
| Luojia Score(mm) | 181.94±75.53 | 117.88±52.48 | 241.30±33.23 | <0.001 |

**Table 2:**
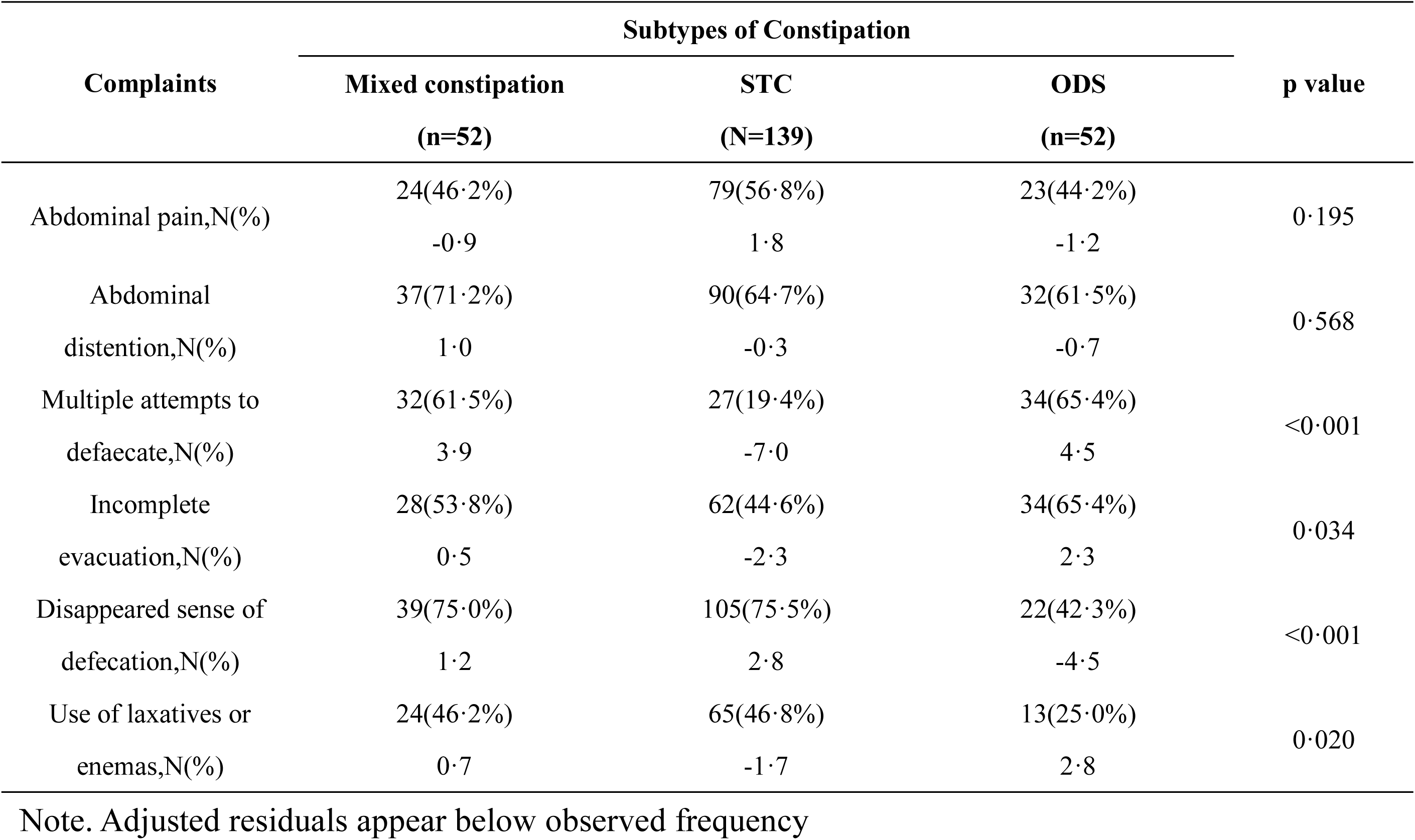
Complaints of patients with constipation

### Quantitative measurement on AXRs

In our study of AXRs, we found that patients with slow colonic transit were more likely to have severe ptosis of transverse colon, which was consistent with other studies (Figure 2). The difference in radiological parameters measured on AXRs, including HI, SI, TI, CS, SH, and Luojia score, between patients with slow or normal colonic transit is shown in Table 1. The HI value is lower in patients with slow colonic transit in comparison with patients with normal colonic transit (81·69±35·28 vs 113·95±43·59 mm, p˂0·001), indicating that they had lower position of the hepatic flexure. There was no statistically significant difference in SI value between the two groups (140·75±35·86 vs 146·57±40·44 mm, p=0·144). The TI value was extremely smaller in patients with slow colonic transit (−100·55±38·52 vs 28·69±72·23 mm, p˂0·001). The CS value was significantly lower in patients with slow colonic transit (22·46±29·79 vs 37·94±32·27 mm, p˂0·001), indicating that the splenic flexure in patients with slow colonic transit was rather closer to the diaphragm. As a result, patients with slow colonic transit had significantly larger SH value (59·06±37·73 vs 32·62±32·15 mm, p˂0·001) and larger Luojia score (241·30±33·23 vs 117·88±52·48, p˂0·001) than those with normal colonic transit.

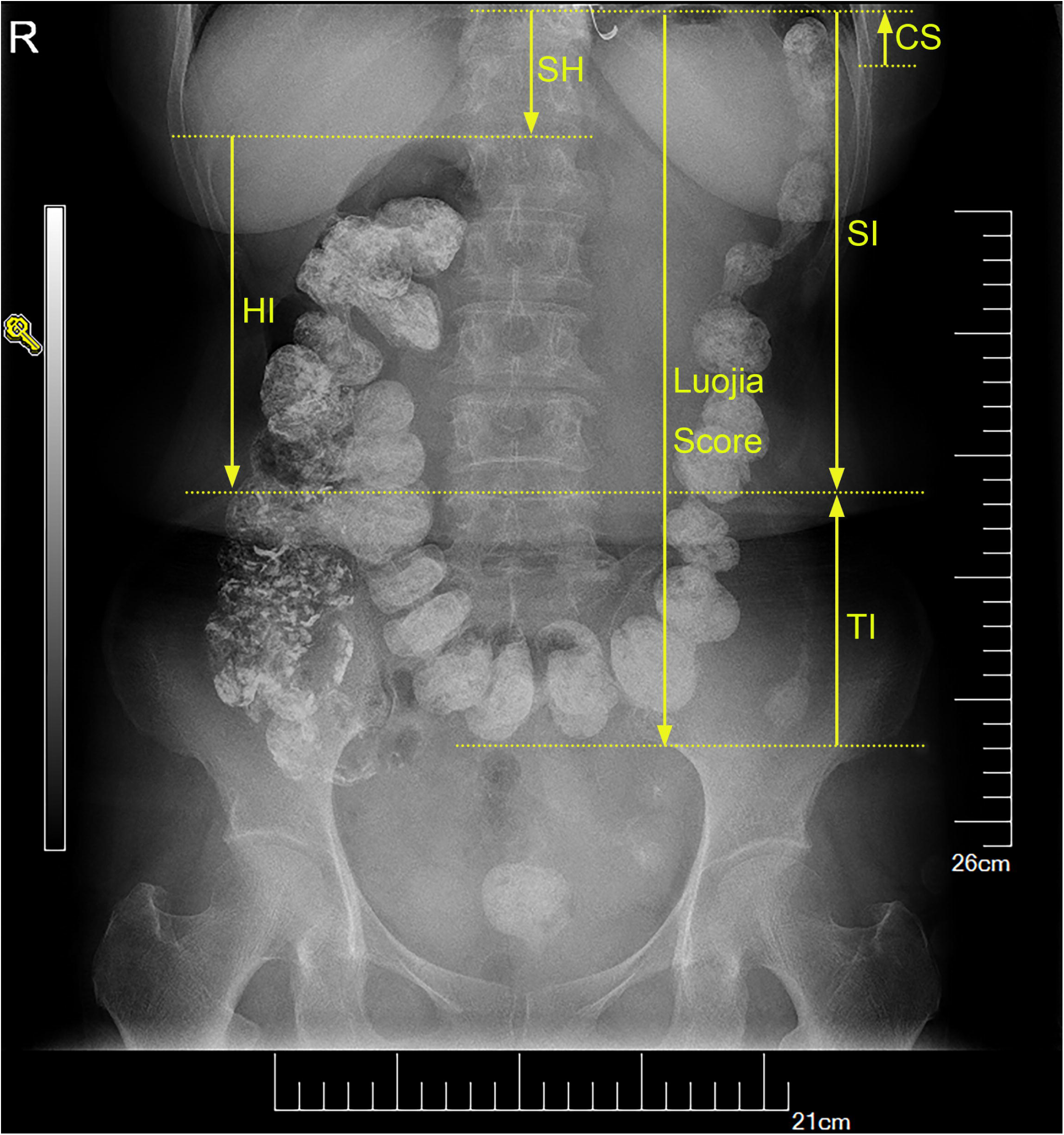

To explore which factors contributed most to the development of slow colonic transit with statistical significance, we analysed the bowel habits and radiological parameters of each group. Significant differences were found in all radiological parameters among subgroups. Results are shown in Table 3 and Figure 3.

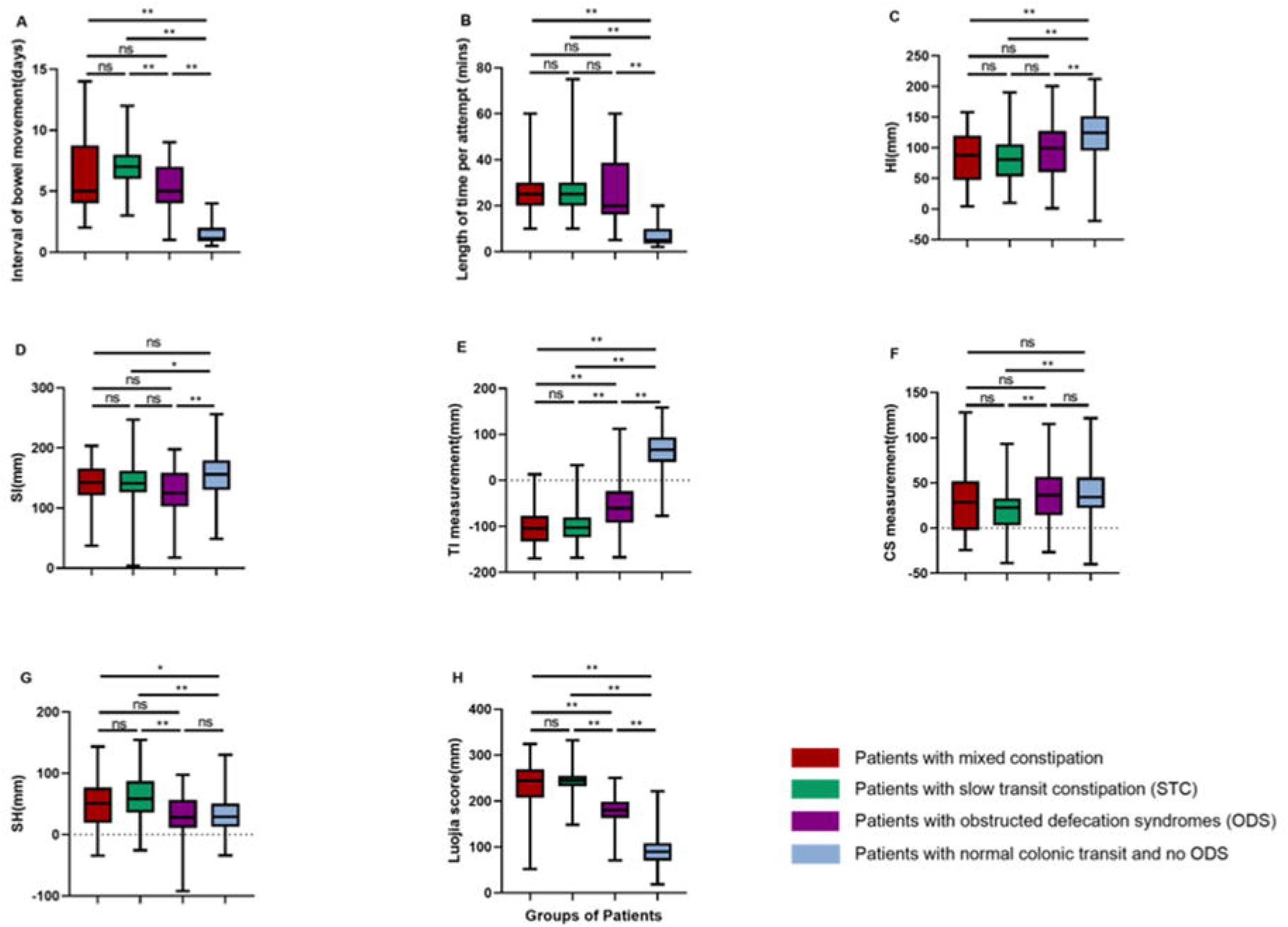

**Table 3:** Bowel habits and radiological parameters on AXRs(n=368). Values are Medium(IQR)

|  | Groups of Subjects |  |  |  | Kruskal-Wallis H test | p value<sup>a</sup> |
| --- | --- | --- | --- | --- | --- | --- |
|  | Mixed constipation<br>(n=52) | STC<br>(n=139) | ODS<br>(n=52) | Normal<br>(n=125) |  |  |
| Interval of bowel movement(days) | 5(4,8·25) | 7(6,8) | 5(4,7) | 1(1,2) | 257·138 | <0·001 |
| Length of time per attempt(mins) | 25(20,30) | 25(20,30) | 20(18·75,36·25) | 5(4,10) | 226·620 | <0·001 |
| HI(mm) | 87·98(47·72,118·46) | 80·84(53·78,105·04) | 99·27(60·78,123·50) | 124·50(97·20,151·40) | 68·752 | <0·001 |
| SI(mm) | 142·63(123·02,164·97) | 141·41(126·87,162·02) | 124·92(104·84,152·85) | 156·00(131·45,179·68) | 19·944 | <0·001 |
| TI(mm) | -104·47(-132·79,-78·96) | -102·83(-123·82,-81·01) | -60·06(-91·74,-23·71) | 66·40(40·82,93·70) | 243·800 | <0·001 |
| CS(mm) | 28·67(0·00,51·55) | 22·67(3·37,32·97) | 36·65(14·80,56·62) | 34·60(22·30,56·20) | 28·924 | <0·001 |
| SH(mm) | 51·03(19·60,75·63) | 58·38(35·80,87·19) | 28·08(11·93,56·21) | 29·30(13·80,50·00) | 49·046 | <0·001 |
| Luojia Score(mm) | 244·27(207·54,268·44) | 244·24(232·25,254·80) | 180·39(164·00,195·95) | 89·40(70·33,108·30) | 265·115 | <0·001 |

a: Results of Kruskal-Wallis H test of all four groups.

We explored the relationship between radiological parameters on AXRs and clinical features of the four subgroups to determine the most appropriate one for evaluating slow colonic transit. As shown in Table 4, Luojia score had an extremely strong association (r=0·834, p˂0·01) with the interval of bowel movements and a moderate association (r=0·542, p˂0·01) with length of time per attempt, which ranked highest among all these radiological parameters. There was also a weak correlation between the Luojia score and BMI (r=-0·275, p˂0·01), indicating that patients with higher Luojia score were likely to be more emaciated.

**Table 4:** Correlations between radiological parameters on AXRs and clinical reports in all participants

|  | HI | SI | TI | CS | SH | Luojia Score |
| --- | --- | --- | --- | --- | --- | --- |
| Age (years) | -0.004 | -0.070 | -0.059 | -0.066 | 0.067 | 0.032 |
| BMI (kg/m<sup>2</sup>) | 0.203** | 0.185** | 0.322** | -0.066 | -0.043 | -0.275** |
| Bowel habits |  |  |  |  |  |  |
| Interval of bowel movement(days) | -0.346** | -0.059 | -0.756** | -0.223** | 0.333** | 0.834** |
| Length of time per attempt(mins) | -0.302** | -0.196** | -0.561** | -0.086 | 0.144** | 0.542** |
| Radiological parameters on AXRs |  |  |  |  |  |  |
| HI (mm) | 1.000 | 0.574** | 0.634** | -0.126* | -0.552** | -0.434** |
| SI (mm) | 0.574** | 1.000 | 0.486** | -0.672** | 0.366** | -0.050 |
| TI (mm) | 0.634** | 0.486** | 1.000 | -0.025 | -0.225** | -0.897** |
| CS (mm) | -0.126* | -0.672** | -0.025 | 1.000 | -0.541** | -0.311** |
| SH (mm) | -0.552** | 0.366** | -0.225** | -0.541** | 1.000 | 0.442** |
| Luojia Score (mm) | -0.434** | -0.050 | -0.897** | -0.311** | 0.442** | 1.000 |

Note. \*: p<0.05; \*\*: p<0.01

### Grading Luojia score

The Luojia score showed a significant difference among the four subgroups. The Luojia score is significantly higher in the slow colonic transit group than that in the normal colonic transit group (241·30±33·23 vs 117·88±52·48 mm, p˂0·001). ROC curve analysis revealed an AUC of 0·964 and 95% confidence interval (CI) of 0·945–0·983 (Figure 4A). The ideal cut-off determined by the Youden’s J statistic, which maximizes specificity and sensitivity, was 195 mm (sensitivity 0·937 and specificity 0·915).

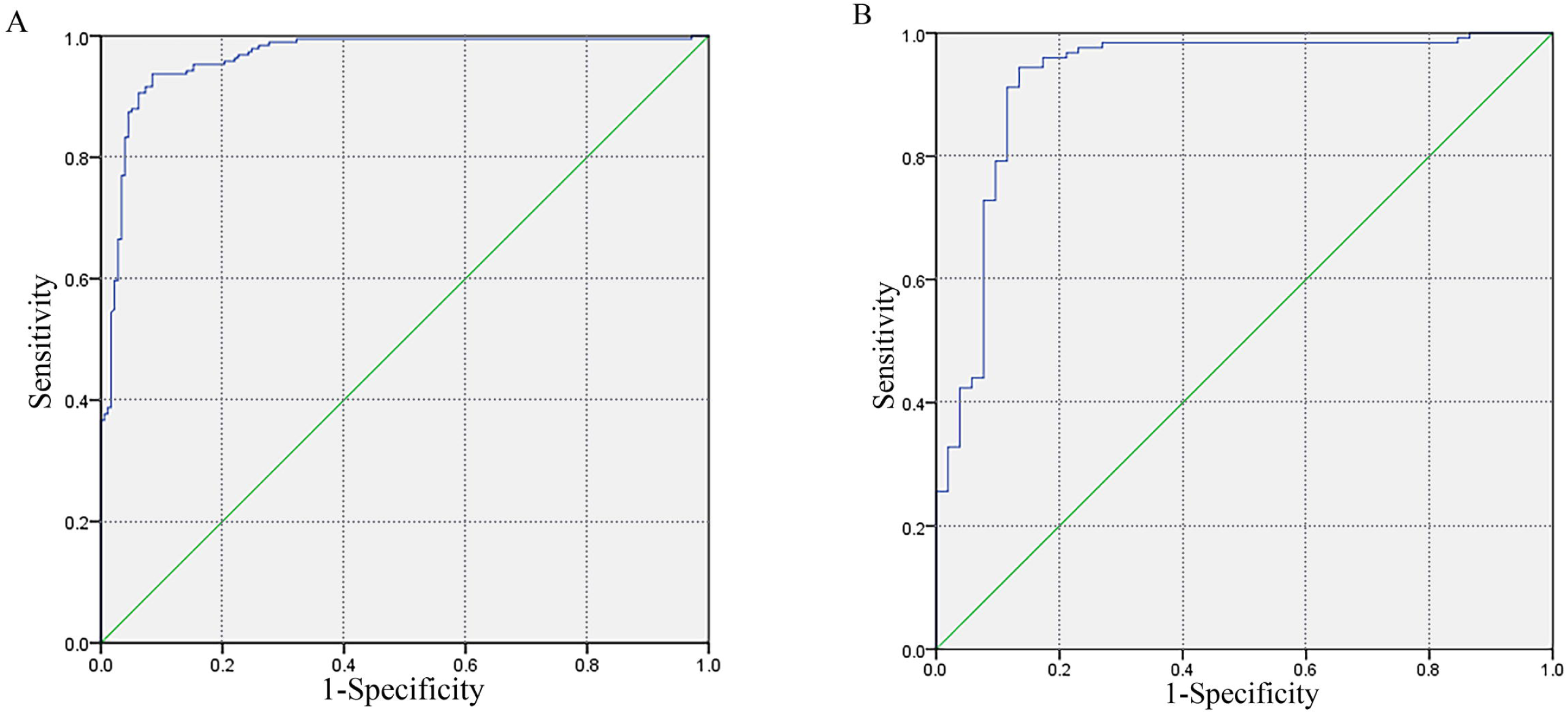

Significant differences in Luojia score were found between the ODS and normal group (177·68±42·00 vs 93·00±32·57 mm, p˂0·001) (Figure 3), and we tried to determine the ideal cut-off to distinguish the ODS and normal group. ROC curve analysis was conducted and revealed an AUC of 0·925 and 95% CI of 0·875–0·976 (Figure 4B). The ideal cut-off was 140 mm (sensitivity 0·865 and specificity 0·944).

### Severity evaluation of STC

Patients with STC were especially required to complete the WCS and HADS for further evaluation of the severity of constipation. The relationship between radiological parameters on AXRs and disease severity of STC is shown in Table 5. Of 139 patients with STC, the average WCS score was 17·51±2·12. As for the evaluation of HADS, 16 patients failed to complete the questionnaires due to resistance to psychological questions. The average HADS score for anxiety and depression were 12·91±2·12 and 12·31±1·87, respectively, in the remaining 123 patients with STC. In patients with STC, the Luojia score was found to have a strong association with WCS (r=0·618, p˂0·001), moderate association with HADS-Anxiety (r=0·507, p˂0·001), and weak association with HADS-Depression (r=0·308, p˂0·001).

**Table 5:**
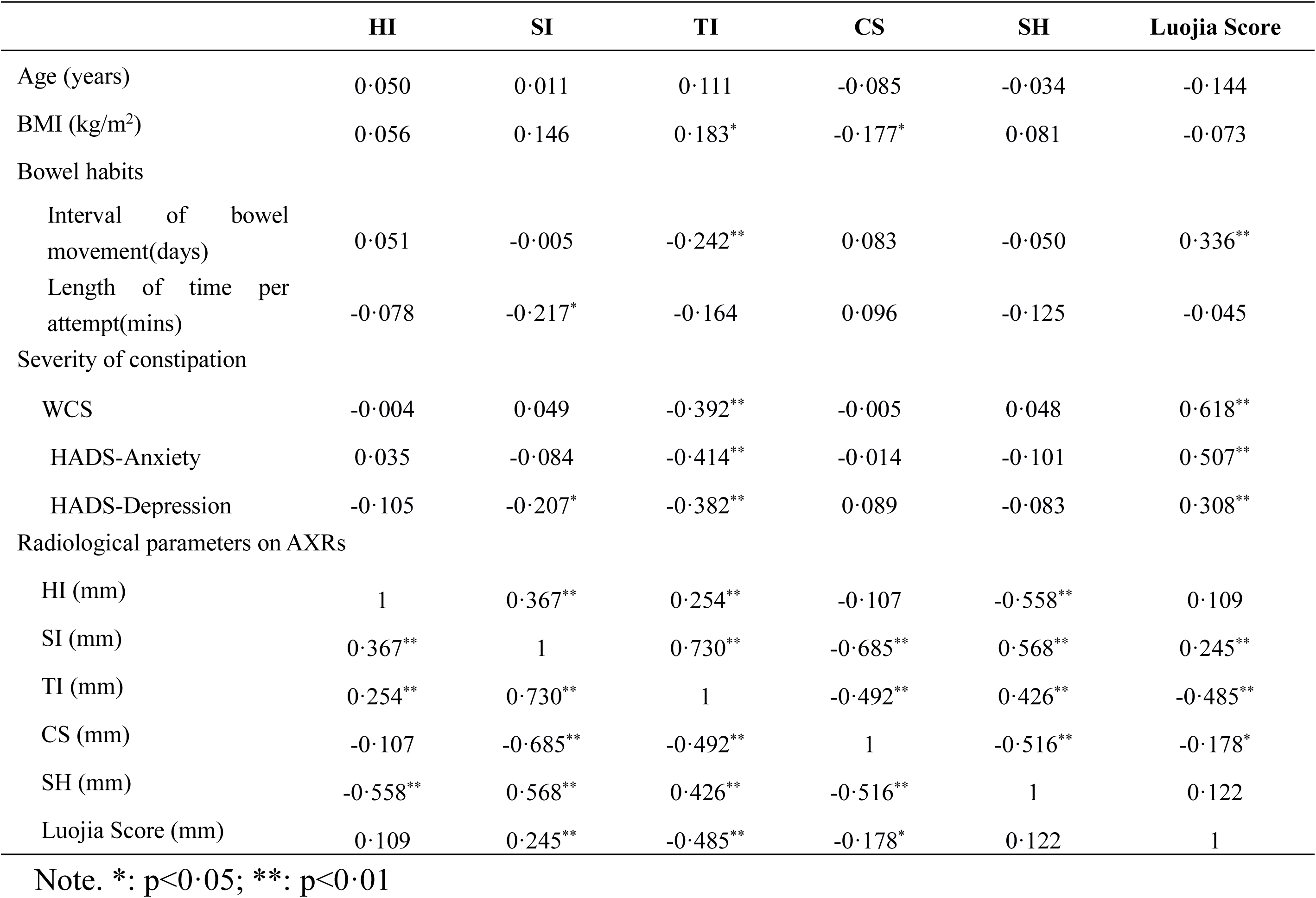
Correlations between radiological parameters on AXRs and disease severity of STC

|  | HI | SI | TI | CS | SH | Luojia Score |
| --- | --- | --- | --- | --- | --- | --- |
| Age (years) | 0.050 | 0.011 | 0.111 | -0.085 | -0.034 | -0.144 |
| BMI (kg/m<sup>2</sup>) | 0.056 | 0.146 | 0.183* | -0.177* | 0.081 | -0.073 |
| Bowel habits |  |  |  |  |  |  |
| Interval of bowel movement(days) | 0.051 | -0.005 | -0.242** | 0.083 | -0.050 | 0.336** |
| Length of time per attempt(mins) | -0.078 | -0.217* | -0.164 | 0.096 | -0.125 | -0.045 |
| Severity of constipation |  |  |  |  |  |  |
| WCS | -0.004 | 0.049 | -0.392** | -0.005 | 0.048 | 0.618** |
| HADS-Anxiety | 0.035 | -0.084 | -0.414** | -0.014 | -0.101 | 0.507** |
| HADS-Depression | -0.105 | -0.207* | -0.382** | 0.089 | -0.083 | 0.308** |
| Radiological parameters on AXRs |  |  |  |  |  |  |
| HI (mm) | 1 | 0.367** | 0.254** | -0.107 | -0.558** | 0.109 |
| SI (mm) | 0.367** | 1 | 0.730** | -0.685** | 0.568** | 0.245** |
| TI (mm) | 0.254** | 0.730** | 1 | -0.492** | 0.426** | -0.485** |
| CS (mm) | -0.107 | -0.685** | -0.492** | 1 | -0.516** | -0.178* |
| SH (mm) | -0.558** | 0.568** | 0.426** | -0.516** | 1 | 0.122 |
| Luojia Score (mm) | 0.109 | 0.245** | -0.485** | -0.178* | 0.122 | 1 |

Note. \*: p<0.05; \*\*: p<0.01

## Discussion

In this retrospective study of 368 patients on the assessment of constipation in a single centre, through AXR, the Luojia score system was graded to evaluate STC based on more severe ptosis of transverse colon in patients with slow colonic transit. Furthermore, we found this system had a good correlation with clinical features.

It was more common to identify anatomy anomaly of transverse colon ptosis on AXRs in patients with colonic constipation.^10-13^ On one hand, patients with slow colonic transit had colonic motor disturbances that might obstruct colonic transit. Since retrograde propulsion, in the opposite direction of the anus, usually occurs in every part of the colon in physiology after a meal and when individuals withhold defecation^20,21^ and in cases with transverse colon ptosis, the transverse colon accounts for 60% of region of interest in AXRs; therefore, severe retention of stool would occur in the transverse colon. The position of the transverse colon becomes lower under the effect of gravity, and this will in turn aggravate the severity of constipation.^11^ On the other hand, transverse colonic ptosis led to relatively elevated splenic flexure, and the increased altitude differences between the transverse colon and splenic flexure increased the static energy needed for faecal evacuation and further aggravated intestinal transmission function.^11^ The Luojia score, a new concept that was defined as the vertical distance from the splenic flexure to the lowest point of transverse colon (SI minus TI) and represented the degree of transverse colon ptosis, was introduced to assess and evaluate whole CTT from plain film AXRs. In our study, an increased Luojia score (241·30±32·23 vs 117·88±52·48) was identified in slow colonic transit group, which presented evident transverse colon ptosis. Moreover, compared to the SI, the increased Luojia score and SH (59·06±37·73 vs 32·62±32·15) suggested the sharp splenic flexure and deep V in the transverse colon.^12^ Briefly, there might be a bidirectional relationship between transverse colon ptosis and constipation, which was found based on the Luojia score.

Precise assessment and evaluation of whole gut and/or colonic transit are important in corroborating or refuting a clinical diagnosis of constipation in patients with diagnostic uncertainty.^22^ To investigate whole CTT, several methods currently exist. Based on plain film AXR, the radiopaque marker method or barium swallow-based method was considered as the traditional approach for colonic transit measurement. Although, with the development of imaging technology, gamma scintigraphy, magnetic tracking system approach, and MRI also have been used in the identification of colonic transit disorder, there is still an unmet demand for a concise and efficient evaluation method for the measurement of colonic transit.^7,23^ David et al. used the Leech score to determine colonic stool burden on AXR as a surrogate for diagnosis of constipation.^7^ Our preliminary data showed that the radiological parameter Luojia score was larger in the slow colonic transit group than that in the normal colonic transit group (241·30±33·23 vs 117·88±52·48, p˂0·001). An ideal cut-off of 195 mm was used in distinguishing the slow transit group and normal colonic transit group with high specificity and sensitivity (sensitivity 0·937 and specificity 0·915). Compared to the Leech score system, which considered visible stool burden as the primary variable of interest, our Luojia score also showed advantages in identifying ODS and normal subgroup with an ideal cut-off of 140 mm (sensitivity 0·865 and specificity 0·944). In this study, we also found that there were significant differences in TI value between the slow colonic transit group and normal transit group and ODS and normal transit group. Among multiple scoring systems for grading stool burden, the Leech method had the highest reproducibility and interobserver agreement.^24^ Although the Leech method defined a score of 7 as a cut-off in delineating slow colonic transit constipation from normal colonic transit constipation, the system showed an unsatisfactory specificity and unneglectable batch effect within two observers. Overall, the Luojia score, an objective measure of assessment defined as the vertical distance from the splenic flexure to the lowest point of transverse colon, might be a sensitive and convenient scoring system in constipation evaluation.

A correlation analysis with clinical characteristics as defecation frequency is helpful in evaluating clinical usefulness of a new scoring system for constipation. In the present study, we explored the relationship between radiological parameters on AXRs and clinical complaints. Moreover, the investigation showed that there was an extremely strong association (r=0·834, p˂0·01) between the Luojia score and interval of bowel movements and moderate association (r=0·542, p˂0·01) between the Luojia score and length of time per attempt. Questionnaires, such as Patient Assessment of Constipation Symptom (PAM-SYM) and WCS, including medical history and symptoms, were the main part of subjective measures of constipation.^25^ We could hardly find correlations between the subjective and objective measures from existing reports. In this study, WCS was used to evaluate the severity of STC. Compared to the poor correlation between the number of ROM and PAM-SYM,^26^ we were satisfied to find that the Luojia score has a strong correlation with WCS (r=0·618, p˂0·001), which indicated that this assessment scale could be used to assess the severity of constipation. Generally, the Luojia score could be considered as a useful surrogate in the evaluation of slow colonic transit.

Although digestive complaints were the most common clinical manifestations of constipation, 65% of patients with constipation had psychological impairment.^27^ A previous study proved that, compared to healthy subjects, patients with constipation were more likely to have anxiety, depression, and social dysfunction.^27,28^ Besides, when HADS was used in the evaluation of psychosomatic disorders, a positive correlation could be found between gastrointestinal dysfunction and HADS scores.^29^ The brain-gut pathway has been considered as bidirectional, and gastrointestinal disorders could result in abnormalities in autonomic nervous that affects the emotional motor system, leading to changes in the hypothalamus-pituitary-adrenal axis and physiological response.^30^ In our study, the Luojia score had a moderate correlation with HADS-Anxiety (r=0·507, p˂0·001) but a weak correlation with HADS-Depression (r=0·308, p˂0·001), which indicated that the Luojia score had a better predictive value for anxiety than depression. Further studies are needed to determine whether the score could be used as an index in the evaluation of constipation severity.

Our study still has several limitations. First, this was a retrospective study, and we cannot exclude the influence of other uncertain confounding factors. Second, the data of this study were from a single medical centre, and our findings may represent more severe cases of constipation that are less generalizable to the population at large. Third, this Luojia score was established based on a small-sample study, and large-sample clinical trials are needed to verify the system. Lastly, due to the small sample size, it was difficult to assess patients with constipation with different grades through the Luojia score, and this could be performed through further clinical trials.

Therefore, based on the difference in presence of transverse colon ptosis in slow colonic transit and normal colonic transit groups, we established a Luojia score system through AXR. The Luojia score was a useful surrogate for constipation identification for its high sensitivity and specificity and has prominent clinical application value due to its high correlation with clinical severity. Still, we need large-sample clinical trials to verify and optimize this system.

## Contributors

All authors contributed to the study design. KS, YH, HX, XR and BL contributed to data acquisition. QQ, WL and CX contributed to data analysis. KS and JH contributed to imaging analysis. KS, CJ and XX wrote the manuscript. CJ was responsible for the overall content as guarantor. All authors approved the final version of the report.

## Data Availability

Data sharing anonymised individual patient data on request or as required by investigators who have been approved by independent review committee about the use of data. Data should not be used beyond the approved proposals A data access agreement signed by requester is needed to gain data. Requests should be sent to the corresponding authors.

## Acknowledgments

This project was funded by grants from the National Natural Science Foundation of China [No. 81570492], the National Natural Science Foundation of China [No. 81500505] and the Clinical Research Special Fund of Wu Jieping Medical Foundation [No. 320.6750.18467].

